# Saliva offers a sensitive, specific and non-invasive alternative to upper respiratory swabs for SARS-CoV-2 diagnosis

**DOI:** 10.1101/2020.07.09.20149534

**Authors:** Rachel L Byrne, Grant A Kay, Konstantina Kontogianni, Lottie Brown, Andrea M Collins, Luis E. Cuevas, Daniela M Ferreira, Alice J Fraser, Gala Garrod, Helen Hill, Stefanie Menzies, Elena Mitsi, Sophie I Owen, Christopher T Williams, Angela Hyder-Wright, Emily R Adams, Ana I Cubas-Atienzar

**Affiliations:** Liverpool School of Tropical Medicine, UK; National Institute for Health Research, UK; Liverpool University Hospitals NHS Foundation Trust, UK

**Author notes:** Corresponding author: Dr Emily Adams, Liverpool School of Tropical Medicine, Pembroke Place, Liverpool L3 5QA.

**Keywords:** SARS-CoV-2, COVID-19, RT-qPCR, swabs, saliva

## Abstract

RT-qPCR utilising upper respiratory swabs are the diagnostic gold standard for SARS-CoV-2 despite reported low sensitivity and limited scale up due to global shortages. Saliva is a non-invasive, equipment independent alternative to swabs.

We collected 145 paired saliva and nasal/throat (NT) swabs at diagnosis (day 0) and repeated on day 2 and day 7 dependent on inpatient care and day 28 for study follow up. Laboratory cultured virus was used to determine the analytical sensitivity of spiked saliva and swabs containing amies preservation media.

Self-collected saliva samples were found to be consistent, and in some cases superior when compared to healthcare worker collected NT swabs from COVID-19 suspected participants. We report for the first time the analytical limit of detection of 10^−2^and 10^0^ pfu/ml for saliva and swabs respectively.

Saliva is a easily self-collected, highly sensitive specimen for the detection of SARS-CoV-2.

## BACKGROUND

Reverse transcription quantitative polymerase chain reaction (RT-qPCR) assays are the reference standard for virologic confirmation of Severe Acute Respiratory Syndrome Coronavirus 2 (SARS-CoV-2), the causative agent of coronavirus disease 2019 (COVID-19) (1). Testing is usually conducted in nasopharyngeal (NP), oropharyngeal (OP) and nasal/throat (NT) swabs (1–3), although viral shedding can be intermittent (4) and multiple swabs are needed to avoid false negative results.

The collection of swab samples also present challenges to attain good quality, as they are inserted into the nasal or oral cavity for at least 10 seconds and often trigger cough and sneezes (5), potentially creating aerosols (6) and decreasing acceptability for repeat sampling. Sampling technique proficiency varies between operators, especially during self-sampling (5) and the process can be uncomfortable, especially in young children (7). It is also unlikely that swabs can accommodate the global need for testing due to shortages in swabs and transport media (8,9)

Saliva is emerging as a non-invasive alternative to upper respiratory swabs. Recent studies have reported saliva has high sensitivity and greater consistency in temporal sampling than other respiratory swabs in transport media (5,10). However, these studies had small sample size (n=25 and n=62 respectively) and did not report specificity or analytical limits of detection (LOD).

Here, we present a description of the analytical sensitivity of saliva in laboratory-infected control samples to establish the LOD and compare paired saliva and combined nasal and throat (NT) swabs from individuals with suspected SARS-CoV-2 participants to conduct a head to head analysis.

## METHODS

### Participant recruitment

This study was embedded into a prospective study evaluating the performance of multiple diagnostic tests for COVID-19 (*Facilitating A SARS Cov-2 Test for Rapid Triage* (FASTER). Adults with signs and symptoms of suspected COVID-19 attending the Royal Liverpool University (RLUH) and Aintree University Hospitals (AUH) in Liverpool, UK were asked to provide specimens within 24 hours of informed consent given on the day of presentation (D0 ±1 day), figure 1 outlines the studies inclusion and exclusion criteria.

**Figure 1:**
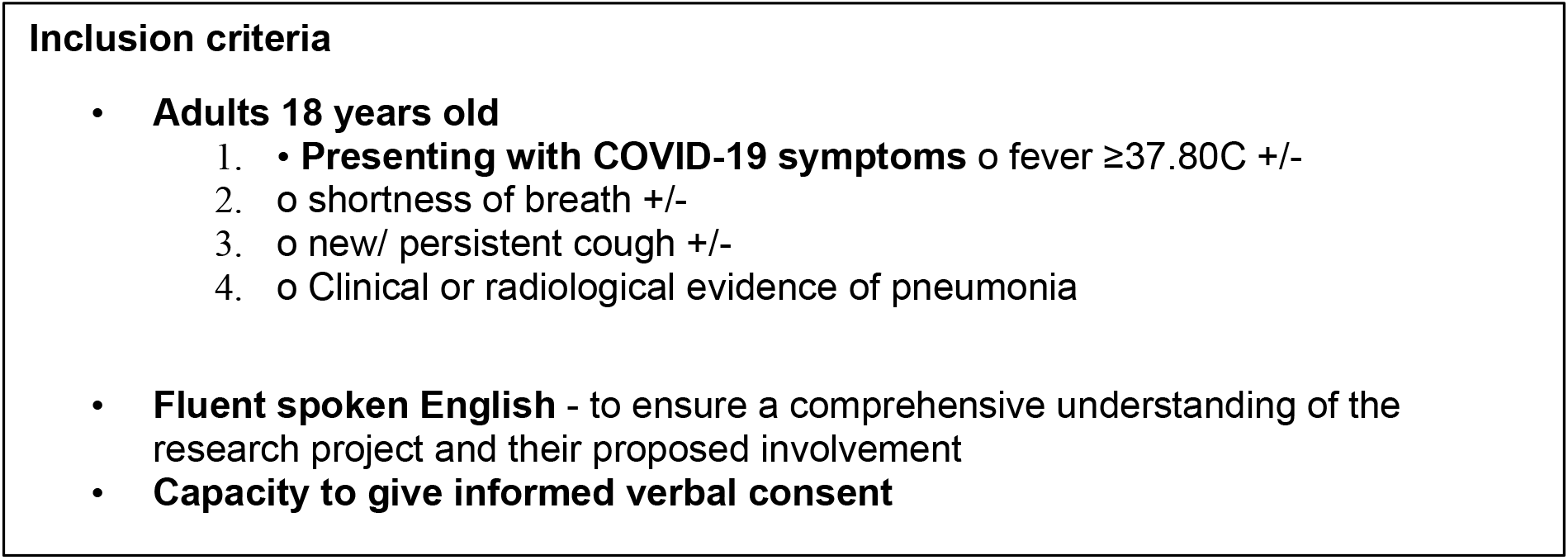
The inclusion and exclusion criteria for the FASTER COVID-19 study.

Further samples were obtained on day 2 ± 1 day (D2) and day 7 ±1 day (D7) dependent on whether participants were positive on D0, continued consent and admitted to the hospital and at day 28 ± 15 days (D28). Ethical approval was obtained from the National Health Service South Central Oxford Committee (20/SC/0169).

### Sample collection

NT swabs (Copan, Italy) were taken by a research nurse delegated to the study. Participants were asked to collect saliva in their mouth and to gently spit it by aid of a funnel in a sterile cryotube collection tube (SARSTEDT, Germany). Saliva volumes were checked by eye and a minimum of approximately 200 microlitres were collected, samples with less volume were excluded from analysis. Samples were transported on ice within 3 hours of collection to the Liverpool School of Tropical Medicine (LSTM) laboratories for processing. RNA extraction was performed immediately on swab samples, while saliva samples were stored at −80°C until processing.

### Analytical sensitivity

Fourteen NT swabs were stored in 1ml of amies preservation medium (Copan, Italy) and 4ml of saliva were collected from a confirmed SARS-CoV-2 negative volunteer. A SARS-CoV-2 isolate (REMRQ0001/human/2020/Liverpool) was propagated in Vero E6 cells (C1008; African green monkey kidney cells), as previously described (11). A serial dilution series of SARS-CoV-2 ranging from 10^6^ to 10^−6^ plaque forming units per ml (pfu/ml) was used to spike 140µl saliva and swab samples. The LOD was determined by the lowest concentration for which all three PCR replicates amplified.

### RNA extraction

Viral RNA was extracted using the QIAamp Viral RNA Mini Kit (Qiagen, Germany) following the manufacturer’s instructions with an internal extraction control incorporated at the lysis stage (Genesig, UK). Non-spiked saliva and NT swab samples from confirmed negative volunteers were included for both matrices as negative extraction controls. Once extracted, samples were taken immediately for downstream application and stored on ice during PCR setup.

### SARS-CoV-2 RT-qPCR

For SARS-CoV-2 RT-qPCR detection, 8µl of extracted RNA was tested using the Genesig® Real-Time Coronavirus COVID-19 PCR assay (Genesig, UK) in a RGQ 6000 thermocycler (Qiagen, Germany). Samples for analytical sensitivity were tested in triplicate, including the negative controls. Samples were classified as RT-qPCR positive if both the internal extraction and the SARS-CoV-2 probes were detected at <40_Ct_. Virus copies/ml were quantified using the manufacturer’s positive control (1.67 × 10^5^ copies/µl) as a reference.

## RESULTS

### Analytical sensitivity

The analytical sensitivity in spiked samples indicated the LOD for saliva was 10^−2^ copies/ml and for the NT swabs 10^0^copies/ml. However, amplification of one or more replicates was recorded for saliva and NT swabs to concentrations 10^−6^ (copies/ml) and 10^−4^ (copies/ml), respectively.

### Participants characteristics

One hundred and ten adults were recruited between April and June 2020. Of these, 61 were female, 49 males. Most participants were hospitalised, with only 21 (19%) discharged home directly from A&E.

In total, 145 paired saliva and NT swabs were collected. Most paired samples were taken at the time of diagnosis (n=110), with follow up samples on D2 (n=14) and D7 (n=6) were only available from positive hospitalised participants. Only 15 participants had returned on D28 at the time of publishing the results.

The proportions of RT-qPCR positive samples for both specimens are shown in Table 1. Overall, 19 saliva and 19 NT swabs (11.6%) of 145 paired samples were positive. As expected, the proportion positive varied with time, with both saliva and NT swabs being more likely to be positive on D2 and D7 than on D28, although numbers are too small to reach statistically significant differences. The viral loads for saliva and NT swabs ranged from 36 to 5.4 × 10^7^ copies/ml for both on enrolment. Overall VLs were similar, with a good agreement between the samples, as shown in Figure 2.

**Tables 1:** Frequency of positive and negative SARS-CoV-2 RT-qPCR in paired saliva and NT swabs from 143 patients with presumptive COVID-19.

|  | Day |  |  |  |  |  |  |  |  |  |  |  |
| --- | --- | --- | --- | --- | --- | --- | --- | --- | --- | --- | --- | --- |
|  | 0 (n=110) |  |  | 2 (n=14) |  |  | 7 (n=6) |  |  | 28 (n=15) |  |  |
|  | Pos | Neg | % (CI95) | Pos | Neg | % (CI95) | Pos | Neg | % (CI95) | Pos | Neg | % (CI95) |
| **NT swabs** | 13 | 97 | 12 (7-20) | 4 | 10 | 29 (8-58) | 2 | 4 | 33 (4-78) | 0 | 15 | 0 (0-22) |
| **Saliva** | 11 | 99 | 11 (5-17) | 5 | 9 | 36 (13-65) | 2 | 4 | 33 (4-78) | 1 | 14 | 5 (1.5-2) |

N = Number, pos = positive, neg = negative, % = percentage positive, CI95 = 95% confidence interval.

**Figure 2:**
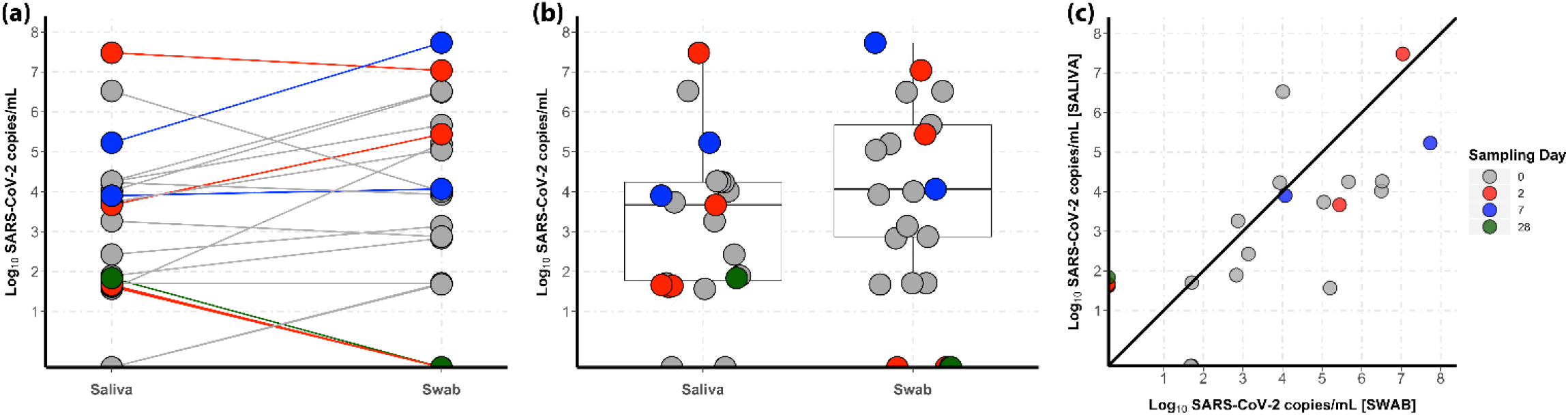
The viral load of RNA recovered from paired saliva and NT swabs on D0 (n=13), D2 (n=5), D7 (n=2) and D28 (n=1). (a) The log viral load (copies/ml) for paired saliva and NT swabs for all days, lines connecting the paired samples. For samples with one specimen as positive and the corresponding pair as negative the data point is plotted but falls below the limit of detection (shown as the grey area) and thus a negative. (b) The mean log viral load (copies/ml) and standard error for paired saliva and NT swabs for all days shown as a box plot with overlaying data points for each sample. (c) The log viral load also depicted as a scatterplot.

The agreement of the RT-qPCR tests is shown in Table 2. Out of 145 paired samples, 23 were positive by at least one sample. Seventeen were positive by both the saliva and NT swabs, 3 (13%) were only positive in saliva and 3 (13%) were only positive in the NT swabs. On D0, 2 samples were RT-qPCR positive in the NT swab but negative in saliva. Nine follow up samples were positive by at least one RT-qPCR. Of these, five were positive in both samples, one was NT swab-positive but saliva negative and 3 were saliva-positive but NT swab-negative.

**Table 2:** Agreement of saliva and nasal/throat swabs SARS-CoV-2 RT-qPCR

|  |  | Nasal/Throat swab |  | All |
| --- | --- | --- | --- | --- |
| Saliva |  | Positive | Negative |  |
| D0 Enrolment | Positive | 12 (%) | 0 (%) | 12 |
|  | Negative | 2 (%) | 96 (%) | 98 |
|  | Total | 14 | 96 | 110 |
| Follow up | Positive | 5 (%) | 3 (%) | 8 |
| D2, D7 + D28 | Negative | 1 (%) | 26 (%) | 27 |
|  | Total | 6 | 29 | 35 |

For one participant, the saliva sample was consistently positive at D0, D2, D7, and D28, whereas the NT swab was positive at day D0, D2 and D7 but negative at D28. All discordant samples that had failed to amplify in saliva or NT swab, the positive sample had a viral load ≤10^1^(copies/ml) and Ct values >36 indicating viral titres were low.

## Discussion

Here we report for the first time the analytical sensitivity of saliva for the detection of SARS-CoV-2 compared to NT swab samples. In spiked saliva samples, SARS-CoV-2 can be detected with greater sensitivity in saliva compared to NT swabs by 100-fold copies/ml, presumably due to the dilution factor of the amies transport medium. We also report the agreement between paired saliva and NT swabs in clinical samples of participants attending hospital settings. Overall, saliva had a good agreement to NT swabs during hospitalisation (D0, D2 and D7) and after recovery (D28). There were minor disagreements between the samples, which were not statistically significant, and these disagreements were bi-directional, with slightly more samples being NT swab positive on enrolment, but more saliva samples being positive during the follow up period. All saliva-positive, NT swab-negative samples in D2 (n=2) and D28 (n=1) reported low viral titre (<10^1^ copies/ml) and are likely due to similar limitations outlined in the analytical sensitivity, a greater dilution factor of amies. It is important to note that these were from participants that had tested positive on D0 and thus likely to be true positive and support saliva to be more consistent for temporal sampling. This is in agreement with Wyllie *et al*., (2020) who reported that saliva had a greater sensitivity and was more likely to be constantly positive throughout the course of infection on a subset of COVID-19 hospitalised participants (n=29).

All saliva-negative but NT swabs-positive were also low viral titre and may have resulted from the different processing time of the two specimens. As NT swabs underwent RNA extraction immediately, saliva samples were stored at −80°C prior to RNA extraction. Freeze-thawing can have a significant impact on the quality of RNA (Ji et al., 2017; Kuang et al., 2018) and it is possible the slightly lower viral loads in saliva were due to this freeze thaw event. Additionally, the lack of preservative media in saliva may reduce the viral detection due to RNA degradation.

Sampling of saliva presents the opportunity to improve our approach to SARS-CoV-2 diagnostics. Saliva is easier to collect; it is non-invasive and much more accepted to the participant than NT swabs and is not reliant on the ability of the participant or staff to take a swab. Moreover, sterile containers are ubiquitous and do not require the purchase of swabs and transport media, which have limited availability during the pandemic. Although saliva is easier to collect, specimens are usually more heterogeneous. This might be due to viral concentrations varying if the participant has coughed recently, cleaned his/her teeth or ingested food or fluids, or the degree of hydration at the time of sampling. This is especially important during acute illness and for patients receiving oxygen therapy. The greatest limitation to our study was the focus on symptomatic hospitalised participants and thereby further data on mild and asymptomatic infection is needed.

It is important to conduct further studies on the effect of storage and transport to document whether RNA and viral loads vary over time without the use of preserving fluids. The use of saliva as a diagnostic sample may be of greatest benefit in low-middle income countries (LMICs) where the pandemic is still accelerating and the availability of swabs for NT sampling are in very low supply. (Gilbert et al., 2020). There will be numerous applications also in the UK for home sampling and the screening of children, who have previously been shown to reject swabbing. As well as in research studies that require repeat sampling and due to the non-invasive nature of saliva will likely promote compliance by participants.

As we move towards the first global peak of the SARS-CoV-2 pandemic it’s imperative that we continue to investigate the optimum diagnostic strategy. Samples that are easy to collect and shown to be effective, such as saliva, warrant the need for immediate validation in certified clinical laboratories.

## Data Availability

Recruitment is ongoing so data isn't currently available. Upon completion we will publish all the findings from the FASTER study.

## Ethical considerations

The study was reviewed and approved by the National Health Service Research Ethics Committee (20/SC/0169) IRAS number 282147.

## Acknowledgements

We are grateful to all the participants recruited to the FASTER clinical research project, for taking the time in this pandemic situation to be involved in research. We also thank the LUHFT and NIHR research nurses and LSTM team who assisted with the sample collection and processing. We thank Ian Pattinson for assistance with viral culture and Ghaith Aljayyoussi for visualising the data in R and. We acknowledge the support of CRN NWC & LHP.

